# Multidrug-Resistant Tuberculosis Rates Among High-Risk Populations in Brazil: An Analysis from 2001 to 2021

**DOI:** 10.64898/2026.11.15.26368155

**Authors:** Isabela Santos de Marins, Carlos Dailton Guedes de Oliveira Moraes, Paulo Otávio Pinto Alves Ferreira, Vitor Augusto Pinto Alves Ferreira, Carlos Alberto Ribeiro Júnior, Hélio Armenara Namorato, João Edimar da Costa Júnior, Fátima Arthuzo Pinto, Alessandro Eustaquio Campos Granato

**Affiliations:** Acadêmicos de Medicina da Faculdade de Ciências Médicas de São José dos Campos - FCMSJC – Humanitas; Docente do Curso de Medicina da Faculdade de Ciências Médicas de São José dos Campos - FCMSJC – Humanitas; Instituto de Ciência e Tecnologia – Universidade Estadual Paulista

**Keywords:** multidrug-resistant tuberculosis, MDR-TB, vulnerable populations, DATASUS, tuberculosis treatment

## Abstract

Multidrug-Resistant Tuberculosis (MDR-TB) represents a global public health threat, with a rising prevalence, particularly among vulnerable populations. This cross-sectional study analyzed MDR-TB notifications in Brazil from 2001 to 2021, using data from DATASUS, and stratified by periods and risk groups (HIV-positive individuals, smokers, drug users, healthcare workers, incarcerated individuals, and people experiencing homelessness). Descriptive analyses and parametric statistical tests (ANOVA and Tukey, p<0.05) were applied. A significant increase in cases was observed across all groups, especially between 2017 and 2021, with growth ranging from 294% to 574% depending on the population. Factors such as treatment abandonment, coinfection, social vulnerability, and occupational exposure were key determinants for MDR-TB dissemination. The findings indicate that current public health policies are insufficient, highlighting the urgent need for targeted prevention strategies, therapeutic follow-up, and control measures for high-risk populations.

## Introduction

Tuberculosis (TB) is a notifiable infectious disease worldwide caused by *Mycobacterium tuberculosis*^1^. It is a life-threatening condition that has afflicted humanity since ancient times, contributing substantially to morbidity and mortality^2^. Currently, the World Health Organization (WHO) classifies TB as a high-risk disease for humanity and considers it one of the leading threats to global public health. The WHO estimates that approximately 10 million people become infected with the disease, with about 6.4 million individuals diagnosed with TB, and that it causes approximately 1.6 million deaths worldwide^3^. According to the United Nations (UN), 95% of cases occur in underdeveloped countries^4^. Brazil has faced persistent challenges related to TB for decades, without achieving effective control of its incidence, prevalence, or treatment outcomes^5^.

Multidrug-resistant tuberculosis (MDR-TB) is defined by resistance to at least rifampicin and isoniazid, whereas extensively drug-resistant tuberculosis (XDR-TB) refers to MDR-TB with additional resistance to at least one fluoroquinolone and one second-line injectable drug^6^. Its resistant form was first documented in 1940, and since then it has posed significant challenges to disease control^7^. The likelihood of developing drug-resistant TB results from a combination of intrinsic and extrinsic factors^8^. Intrinsic pathogen-related aggravating factors include a vast reservoir of latently infected populations, challenges in diagnosis and case tracking, prolonged treatment duration associated with multiple adverse effects, and treatment interruption before completion of the therapeutic regimen. Other major concerns involve extrinsic factors, such as HIV coinfection, low socioeconomic status, and limited access to healthcare services. These characteristics contribute to the persistence and spread of drug-resistant TB^6,9^. Isoniazid and rifampicin are considered the cornerstone drugs of tuberculosis treatment; however, in the drug-resistant form of the disease, they are insufficient, thereby rendering the therapeutic regimen more complex. Consequently, clinicians must rely on alternative drug combinations, often associated with more severe adverse effects and prolonged treatment duration^10^.

In 2012, approximately 45,000 cases of MDR-TB were identified, with an estimated 170,000 deaths reported^2^. By 2016, an estimated 490,000 new cases of MDR-TB had occurred, making the scenario particularly concerning due to the prospect of accelerated growth^7^. Among all infectious diseases, tuberculosis has the highest mortality rate worldwide, exceeding that of HIV/AIDS^11^. Among individuals affected by drug-resistant TB, the most vulnerable groups include people experiencing homelessness, individuals with substance use disorders, persons deprived of liberty, individuals living with HIV or those who are immunocompromised, and healthcare workers^6^. Within this context, the present study aims to analyze the relationship between MDR-TB and the most vulnerable and high-risk populations in Brazil.

### Methodology

This study was designed as a cross-sectional epidemiological investigation, based on data collected from the Departamento de Informática do Sistema Único de Saúde (DATASUS). DATASUS is a platform maintained with data provided by the Ministério da Saúde (Ministry of Health) and is publicly accessible through its official website (http://www.datasus.gov.br)^12^. The information available on this platform is in the public domain; therefore, ethical approval was not required for this study, as it complies with the current principles established by Resolution No. 466/2012 of the Conselho Nacional de Saúde (National Health Council). Furthermore, the dataset contains no personal identifiers, including only information related to public health surveillance^12–15^.

Data collection was conducted through access to the Departamento de Informática do Sistema Único de Saúde (DATASUS) system between September and November 2023. The target population of this study comprised all reported new cases and deaths due to drug-resistant tuberculosis recorded in Brazil between 2001 and 2021. The study also examined the epidemiological profile of the affected population and vulnerable groups, including people experiencing homelessness, individuals with substance use disorders, persons deprived of liberty, individuals living with HIV or those who are immunocompromised, and healthcare workers.

Thus, the study was designed as a population-based investigation using descriptive statistical analysis to characterize demographic and treatment-related variables, including tuberculosis-related complications in Brazil. Temporal analyses were performed over the period from 2001 to 2021, with data analyzed in their entirety. Additionally, the study period was subdivided into four intervals—first period (2001– 2006), second period (2007–2011), third period (2012–2016), and fourth period (2017– 2021)—to assess changes in the epidemiological profile of the disease over time. All analyzed data were presented as means and standard deviations, adopting a significance threshold of p< 0.05.

### Statistical Analysis

Data were tabulated using Microsoft Excel (Microsoft Office Professional Plus, 2016; Microsoft). Statistical analyses were performed using GraphPad Prism version Graphical data presentation was generated using GraphPad Prism 6.0 and the Departamento de Informática do Sistema Único de Saúde (DATASUS) platform (2023). Normality was assessed for all variables using the D’Agostino–Pearson test or the Shapiro–Wilk test. Parametric tests were applied for group comparisons (one-way ANOVA), with confirmation by Tukey’s post hoc test. A significance level of *p* < 0.05 was adopted. All data and graphical outputs were analyzed and presented as means and standard deviations.

## Results

Between 2001 and 2021, a total of 1,620 cases of multidrug-resistant tuberculosis were reported among patients living with HIV in Brazil. The lowest mean number of cases was observed during the period from 2001 to 2006, whereas the highest mean was reported between 2017 and 2021 (1008.00 ± 97.09 cases). Notably, the last evaluated period showed a 574% increase compared with the beginning of the time series. The periods from 2012 to 2016 and from 2017 to 2021 demonstrated a statistically significant difference (*p* < 0.0001) compared with the other periods.

Between 2001 and 2021, a total of 1,620 cases of multidrug-resistant tuberculosis were reported in Brazil. The lowest mean number of cases was observed during the period from 2001 to 2006 (3.2 ± 5.60 cases), whereas the highest mean was reported between 2017 and 2021 (150.4 ± 19.76 cases). During the period from 2007 to 2011, a mean of 27.4 ± 16.65 cases was observed, and between 2016 and 2021, a mean of 144.00 ± 39.73 cases was recorded. The periods from 2012 to 2016 and from 2017 to 2021 showed a statistically significant difference (*p* < 0.0001) compared with the other periods.

Between 2001 and 2021, a total of 7,554 cases of multidrug-resistant tuberculosis were reported in Brazil. The lowest mean number of cases was observed during the period from 2001 to 2006 (2.00 ± 1.86 cases), whereas the highest mean was reported between 2017 and 2021 (1048.20 ± 146.73 cases). Notably, the last evaluated period showed a 294% increase compared with the period from 2012 to 2016. The period from 2017 to 2021 demonstrated a statistically significant difference (*p* < 0.0001) compared with the other periods.

Between 2001 and 2021, a total of 5,217 cases of multidrug-resistant tuberculosis were reported in Brazil. Cases reported during the period from 2017 to 2021 accounted for 98.58% of the total. The period from 2017 to 2021 showed a statistically significant difference (*p* < 0.0001) compared with the other periods.

Between 2001 and 2021, a total of 5,675 cases of multidrug-resistant tuberculosis were reported in Brazil. Cases reported during the period from 2017 to 2021 accounted for 82.18% of the total. The period from 2017 to 2021 showed a statistically significant difference (*p* < 0.0001) compared with the other periods.

Between 2001 and 2021, a total of 6,166 cases of multidrug-resistant tuberculosis were reported in Brazil. Cases reported during the period from 2017 to 2021 accounted for 89.60% of the total. The period from 2017 to 2021 showed a statistically significant difference (*p* < 0.0001) compared with the other periods.

Between 2001 and 2021, a total of 6,670 cases of multidrug-resistant tuberculosis were reported in Brazil. Notably, the last evaluated period showed a 326% increase compared with the period from 2012 to 2016. The period from 2017 to 2021 demonstrated a statistically significant difference (*p* < 0.0001) compared with the other periods. Furthermore, cases reported during the period from 2017 to 2021 accounted for 71.04% of the total.

## Discussion

Multidrug-resistant tuberculosis (MDR-TB) is considered a global health problem, with a prevalence of 11.6%^16^. Over the past two decades, Brazil has experienced a significant increase in the number of cases, as shown in Figure 1. The rate of resistance to at least one drug between 2003 and 2006 was 12.4%^2^. This increase is strongly associated with multiple risk factors and the country’s socioeconomic conditions^7^. Individuals who live or work in environments with a high bacillary load, poor ventilation, and overcrowding are more susceptible to acquiring the disease^17^.

**Figure 1:**
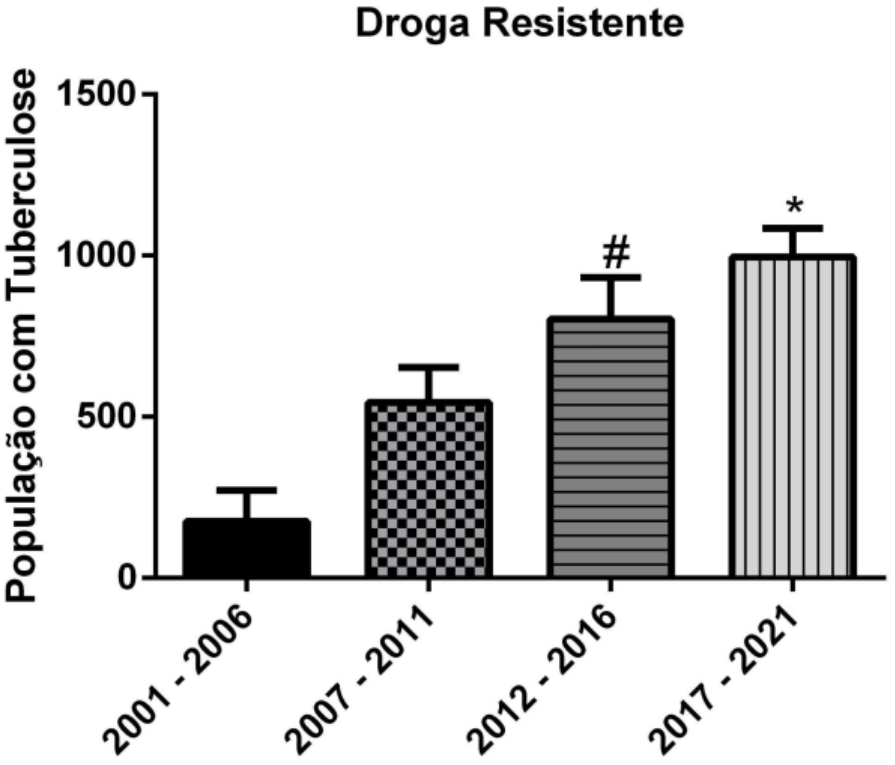
Total Number of Multidrug-Resistant Tuberculosis Cases in Brazil from 2001 to 2021. Source: Authors’ own elaboration

An important property of drug-resistant tuberculosis (DR-TB), compared with drug-susceptible TB, is its greater transmissibility, which represents an increased risk of dissemination and a major concern for public health authorities^18^. Drug-resistant TB may develop as a result of deficiencies in treatment adherence, including poor compliance, treatment abandonment, inappropriate treatment regimen selection, or inadequate drug supply. The management of MDR-TB cases is challenging, particularly in the presence of cavitary lesions and caseous granulomas that harbor subpopulations of bacilli, some exhibiting phenotypic resistance despite genetic susceptibility (non-heritable resistance that regresses after antibiotic withdrawal), while others present genetic mutations associated with both phenotypic and heritable resistance^43^.

A general analysis of the present study’s findings indicates a persistent increase in the incidence and prevalence of MDR-TB cases over time, as illustrated in Figure 1. This increase occurs particularly among populations considered vulnerable, including persons deprived of liberty, people experiencing homelessness, individuals with substance use disorders, smokers, and people living with HIV/AIDS.

A study conducted by Ridolfi and colleagues analyzed the biological and behavioral factors associated with each component of unfavorable tuberculosis treatment outcomes. Among the total number of participants, 13% did not achieve successful treatment outcomes. Among these individuals, a common and prominent characteristic was being people living with HIV. Furthermore, the study identified that behavioral factors, such as current drug use and tobacco consumption, were more strongly associated with loss to follow-up, whereas clinical comorbidities, such as HIV and diabetes, were more closely associated with treatment failure and death. Therefore, intervention strategies focused on health education, support for substance use treatment, and management of comorbidities may be particularly effective in reducing these unfavorable outcomes^20^.

A study by Coelho and colleagues confirmed this hypothesis, demonstrating a significant increase in MDR-TB cases among patients living with HIV, with approximately 50% of patients diagnosed with drug-resistant TB being HIV-positive. These findings are consistent with the data presented in Figure 2, which showed a significant increase in the number of cases^21^. Additionally, these patients presented with a higher frequency of cavitary pulmonary lesions and more frequent changes in treatment regimens. The increase in MDR-TB among patients living with HIV or AIDS is not solely related to immune system impairment, but also to the fact that these individuals are often already receiving multiple long-term medications with substantial adverse effects. This circumstance may hinder adherence to additional daily medications that must be incorporated into their routine for prolonged periods during tuberculosis treatment^22^.

**Figure 2:**
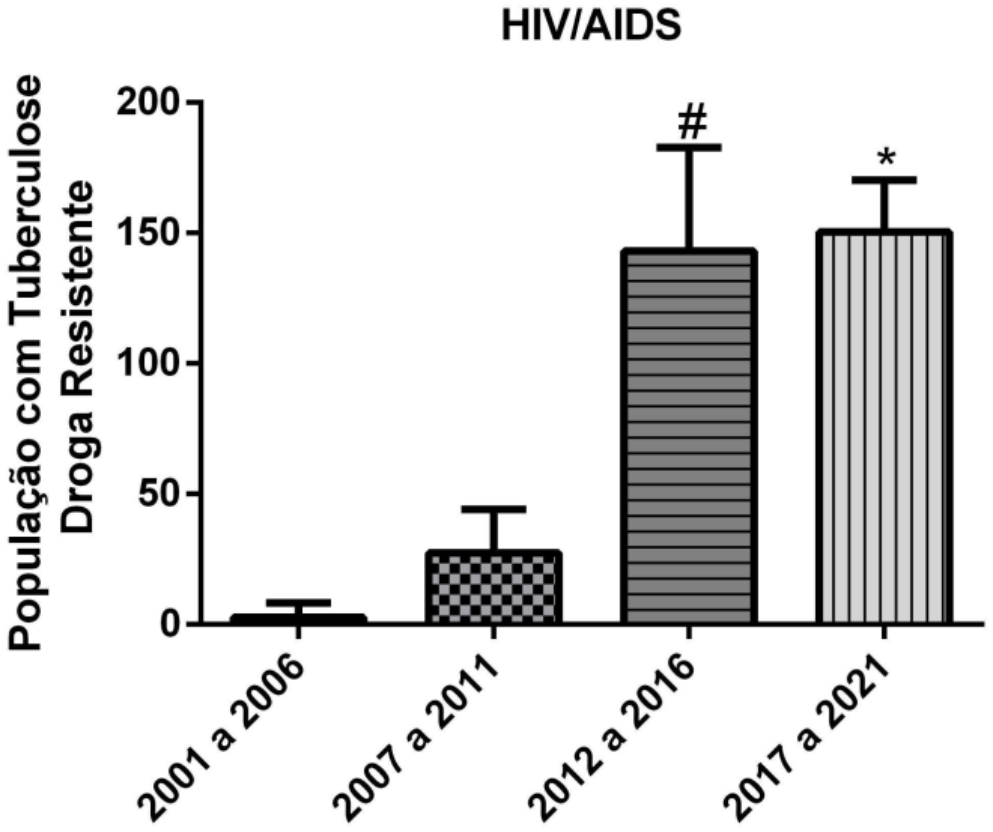
Total Number of Multidrug-Resistant Tuberculosis Cases Among People Living with HIV in Brazil from 2001 to 2021. Source: Authors’ own elaboration

Since 1918, smoking has been identified as a risk factor for tuberculosis, with exposure to smoke being substantially associated with TB recurrence and increased mortality. Studies indicate that more than 1.3 billion smokers live in underdeveloped or developing countries, where tuberculosis rates are also higher^23^. These findings are consistent with the data presented in Figure 3, which demonstrates an increase in the number of cases over time. This association may be explained by the fact that smoking alters the defense mechanisms of the bronchial tree and reduces blood oxygen concentration, contributing to the severity of necrotizing lesions and delaying tissue repair, factors that may lead to the development of more severe disease manifestations. Cigarette smoke plays a role in disease pathogenesis by promoting ciliary dysfunction and reducing immune responses through impaired macrophage activity, thereby increasing susceptibility to mycobacterial infection^24^. Smoking increases the risk of latent and active TB, false-negative results and delays in sputum testing, reduces treatment adherence, and increases recurrence rates; collectively, these factors contribute to the development of multidrug-resistant disease^25^.

**Figure 3:**
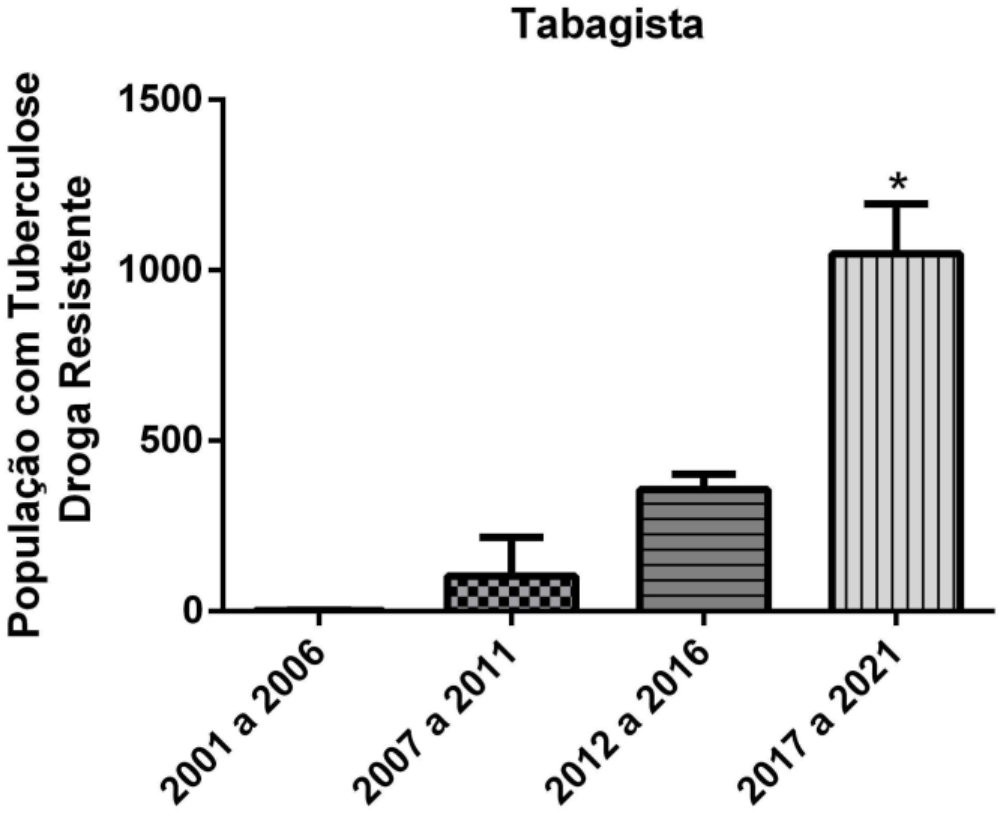
Total Number of Multidrug-Resistant Tuberculosis Cases Among Smokers in Brazil from 2001 to 2021. Source: Authors’ own elaboration

Although healthcare workers are not considered a neglected population, they remain a high-risk group for MDR-TB, as their occupational environment involves frequent contact with bacilliferous patients, including those who do not adhere adequately to treatment. This risk is reflected in the annual tuberculin skin test conversion rate, which ranges from 2.2% to 5.6% ^26^. Such tuberculin conversion is primarily attributed to nosocomial exposure to patients with pulmonary TB, the absence or inadequacy of individual or collective biosafety measures, and is particularly notable among nursing professionals^27^. In this context, tuberculosis is classified as a notifiable occupational disease. An aggravating factor is that these workers, in addition to exposure to bacilliferous patients, are also exposed to other infectious conditions affecting patients. According to the manual of recommendations for tuberculosis control, nursing professionals present the highest risk among healthcare workers, with rates 3 to 20 times higher than those of the general population, followed by clinical pathologists (6–11 times), laboratory technicians (2–9 times), and pulmonologists (6 times) ^28^. The data presented in this study demonstrate a significant increase in MDR-TB notifications among healthcare workers during the period from 2017 to 2021. This finding may be explained by underreporting in previous years, as well as by the lack of effective interconnection between health information systems^20^.

The prison population is of particular concern to public health authorities, as it represents a group characterized by multiple vulnerabilities, including HIV infection, alcohol consumption, and the use of illicit drugs and tobacco, with a growing trend in substance use^29^. Furthermore, poor living conditions in correctional facilities, such as overcrowded shared cells, inadequate ventilation, limited exposure to sunlight, and insufficient healthcare services, predispose individuals to the transmission of TB and its drug-resistant forms^50^. These findings are consistent with the data presented in Figure 5, which demonstrates an increase in MDR-TB rates within this population. Additionally, alcoholism and AIDS have also been identified as important risk factors, as they may weaken the immune system and hinder adherence to TB treatment^31^. Another critical factor identified was a history of previous TB treatment among persons deprived of liberty. Individuals who have undergone prior treatment have an increased risk of developing drug-resistant TB, particularly when previous treatment was not completed adequately or when interruptions occurred in the therapeutic regimen^32^.

**Figure 4:**
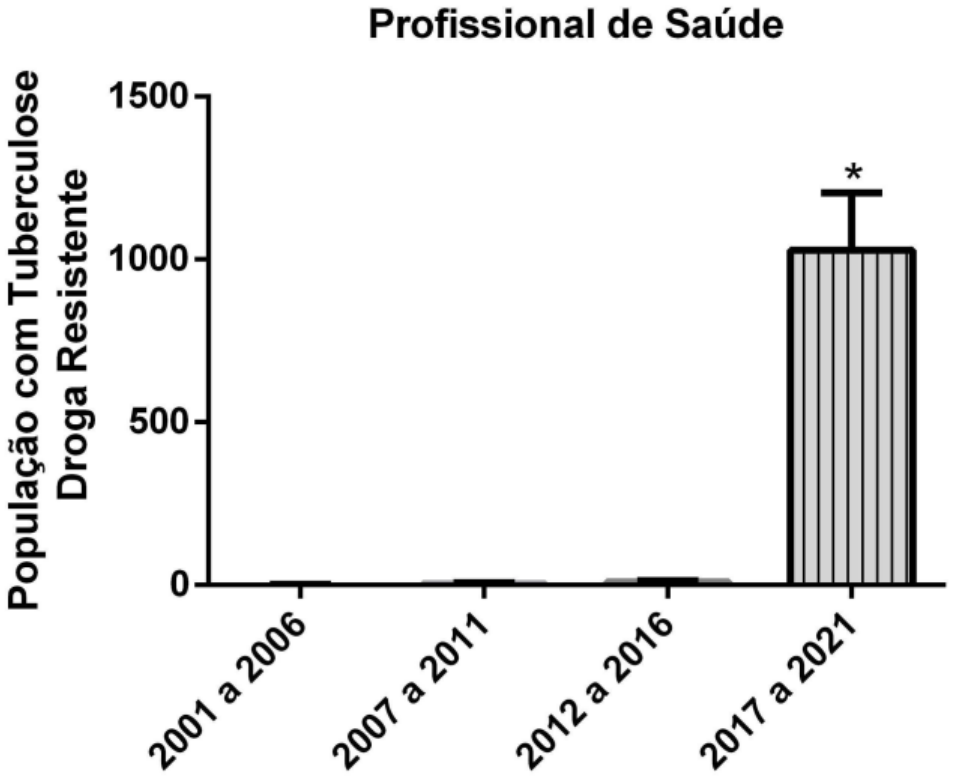
Total Number of Multidrug-Resistant Tuberculosis Cases Among Healthcare Workers in Brazil from 2001 to 2021. Source: Authors’ own elaboration

**Figure 5:**
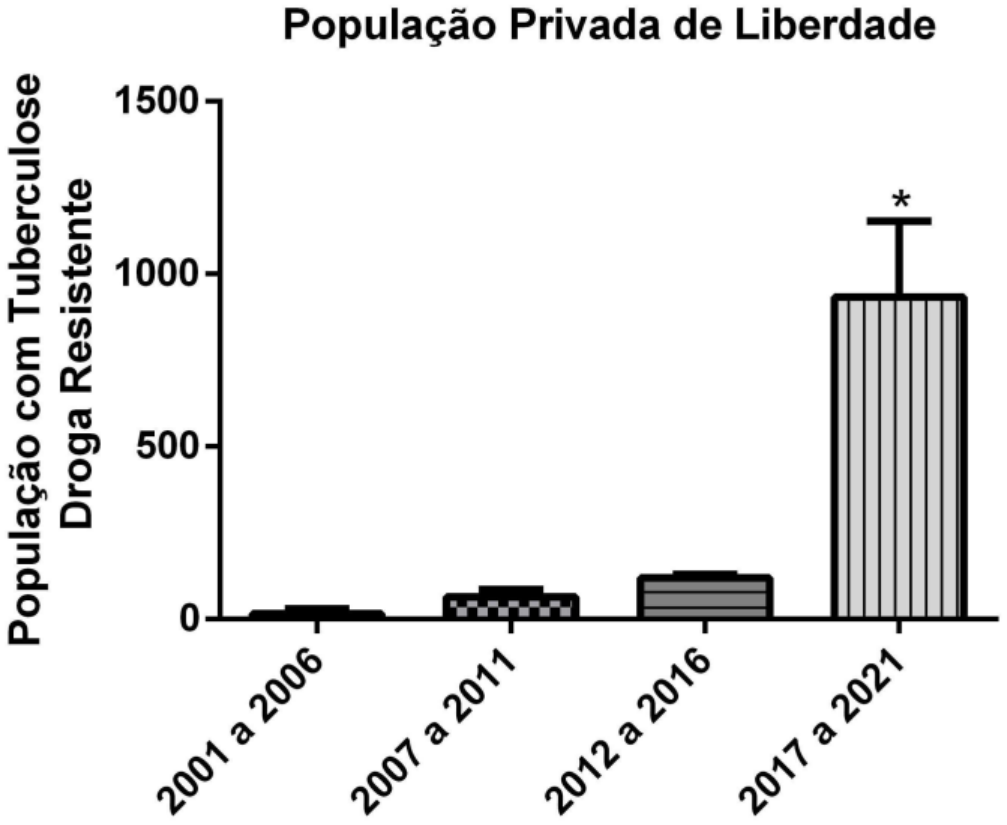
Total Number of Multidrug-Resistant Tuberculosis Cases Among the Prison Population in Brazil from 2001 to 2021. Source: Authors’ own elaboration

Individuals experiencing homelessness are 56 times more likely to be affected by tuberculosis, as they are exposed to multiple vulnerabilities^33^. This finding is consistent with the data presented in Figure 6, which demonstrates an increase in MDR-TB rates within this population. Due to limited financial resources, this group makes less frequent use of healthcare services, resulting in delayed diagnosis, which may lead to poor treatment adherence and unfavorable outcomes^34^. In this context, progression to drug-resistant forms of the disease becomes more likely. Individuals affected by TB tend to experience unsuccessful treatment outcomes due to factors such as intravenous drug use, alcohol consumption, malnutrition, and HIV coinfection. This association is supported by the findings of the present study, which identified a positive intrinsic relationship between high-risk populations and MDR-TB^35^. Furthermore, for people experiencing homelessness, concerns such as safety, access to food, and the need for rest often compete with attention to healthcare needs^36^.

**Figure 6:**
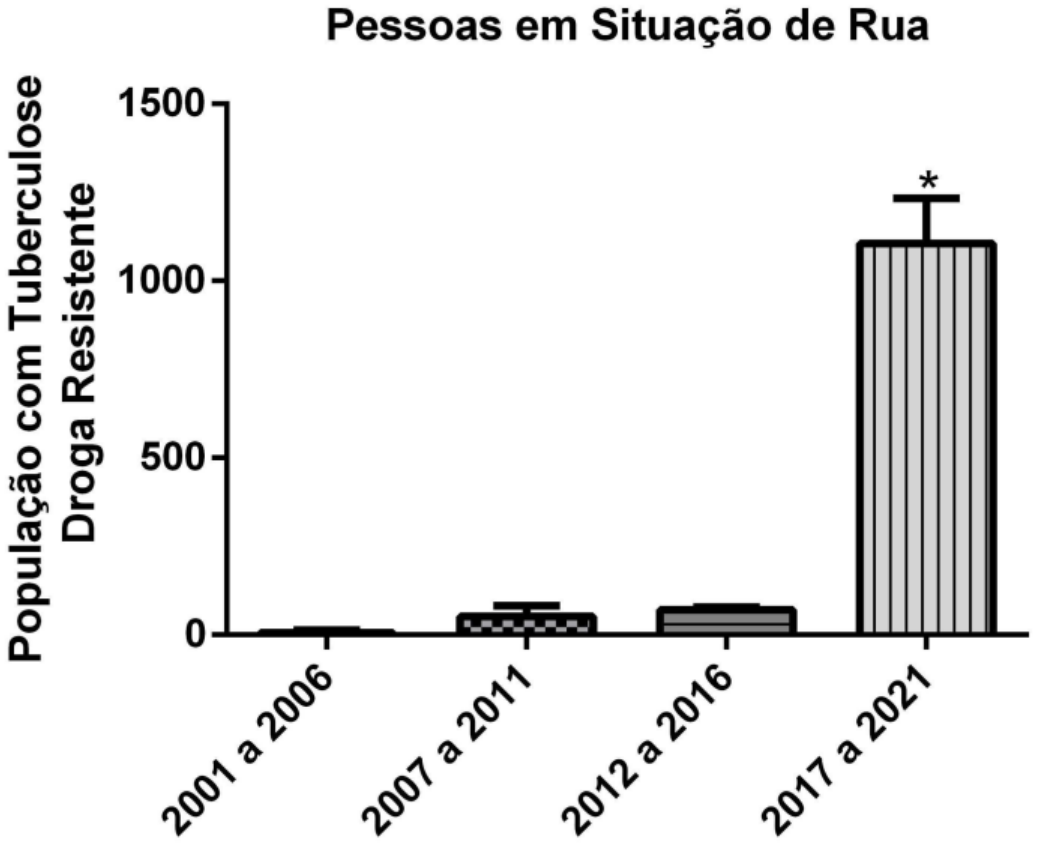
Total Number of Multidrug-Resistant Tuberculosis Cases Among People Experiencing Homelessness in Brazil from 2001 to 2021. Source: Authors’ own elaboration

Individuals with substance use disorders constitute one of the socially marginalized groups requiring special attention in healthcare, as they accumulate multiple risk factors beyond illicit drug use, including a higher likelihood of experiencing homelessness, acquiring infectious diseases through needle sharing, and concurrent dependence on licit substances such as tobacco and alcohol^37^. These findings are consistent with the data presented in Figure 7, which illustrates an increase in MDR-TB rates within this population. A study conducted by Kazempour and colleagues demonstrated that 65.7% of patients with drug-resistant tuberculosis who died were male and smokers, and 50% were drug users^37^. It is noteworthy that estimates indicate that more than 15% of deaths associated with tuberculosis could be prevented if patients did not engage in heavy alcohol consumption^38^. Data from another study suggest that individuals with alcohol use disorder are nearly four times more likely to discontinue tuberculosis treatment, with the main reasons for treatment abandonment being a perceived sense of well-being and adverse effects resulting from the combined use of medications with alcohol and other drugs^39^. Furthermore, even when access to healthcare services is facilitated, adherence to long-term treatment among individuals with substance use disorders remains a significant challenge^38^.

**Figure 7:**
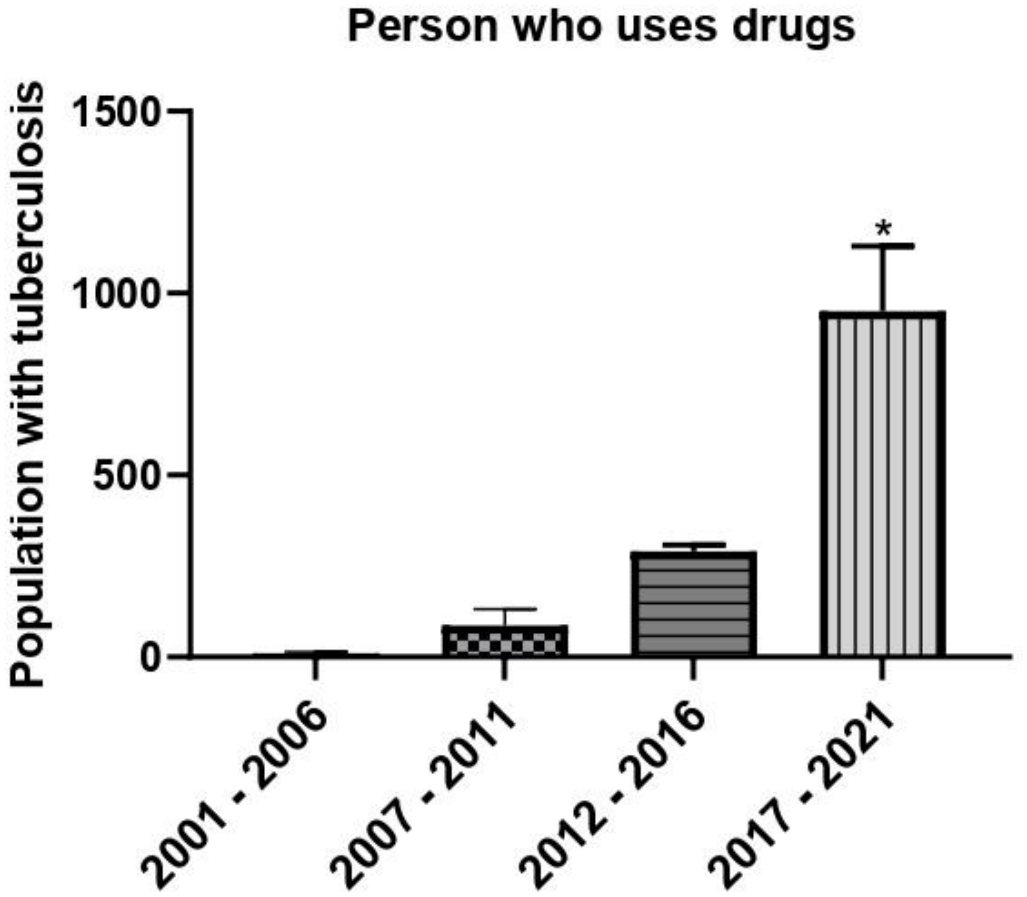
Total Number of Multidrug-Resistant Tuberculosis Cases Among Individuals with Substance Use Disorders in Brazil from 2001 to 2021. Source: Authors’ own elaboration

An analysis of all the figures in the present study indicates that the period from 2001 to 2011 showed the lowest MDR-TB rates, which may be explained by the existence of multiple health information systems that were not integrated at the time. This scenario changed in 2013 with the establishment of the Departamento de Informática do Sistema Único de Saúde (DATASUS), which unified the country’s compulsory disease notification information system^39^.

## Conclusion

Based on the data analysis, it can be concluded that in recent years there has been an overall increase in the number of MDR-TB cases across all high-risk populations studied in Brazil. This increase highlights the limitations of current public health policies aimed at controlling this disease. The higher prevalence of MDR-TB may be attributed to treatment abandonment, poor socioeconomic conditions, and inadequate patient follow-up within the healthcare system. In this context, the findings underscore the urgent need to strengthen public health policies directed at combating this condition.

The main limitations identified in conducting this study included the initial absence of a centralized database compiling all notifications and, even after the establishment of the Departamento de Informática do Sistema Único de Saúde (DATASUS), the incomplete migration of these data to the new platform. Additional limitations included underreporting of cases among healthcare workers prior to 2017 and the inability to perform internal data linkage within the system.

## Data Availability

All content in this article may be considered public; in other words, once submitted, it will be accessible to everyone.

